# Comparative analysis of antibody production by mRNA 1273, AZD 1222 and BBIBP-CorV on elderly people suffering from different co-morbidity in Bangladesh

**DOI:** 10.1101/2021.11.15.21266338

**Authors:** Ashraful Hoque, Md Marufur Rahman, Shahnewaz Parvez, Hossain Imam, Nurun Nahar, Forhad Uddin Hasan Chowdhury

## Abstract

**Background:** As the pandemic spread so quickly all over the world, the scientist did not get the right time to cover all age group populations for a vaccine trial. The elderly population is usually vulnerable for COVID-19 which was proven by different research work and focus was to save this group of people was the prime concern for every country of the world. Though vaccines that got emergency use authorization have proven their efficacy which one is better for elderly people suffering from different co-morbid conditions is still not established. In this study, we want to evaluate the antibody production by mRNA 1273, AZD 1222, and BBIBP-CorV in elderly people with different comorbidity.

**Method:** We include 40 people in each group who have at least one comorbid condition and the total sample size was 120. The sample was taken from them before vaccination and 14 days after the second dose. Adverse event following immunization was recorded if any. Antibody measurement was done by ELISA method by using DiaSino SARS-CoV-2 S1 RBD IgG Quant.

**Result:** Among 120 participants with an equal number of participants in each of the vaccine groups all of them were aged between 60-72 years, of whom 65% were males and 35 % were females. Anti S1 RBD IgG was detected among all the participants from each vaccine group after 14 days of taking their 2nd dose. A non-parametric multiple comparison test (Kruskal-Wallis test) of Anti S1 RBD IgG levels among three vaccine groups revealed significant differences (P-value <0.05) between groups. The IgG level was almost twice in the mRNA-1273 group (mean 577.1± 44.33 AU/ml) compared to AZD1222(mean 308.5 ±37.91 AU/ml) and BBIBP-CorV group.

**Conclusion:** From this small sample size, we predicate that mRNA 1273 produce much higher anti-S1 RBD IgG than the other two vaccines. Every vaccine is safe and effective whose is approved by WHO. Calculative use of the vaccine may produce an outcome for the future as we are still way behind in the proper amount of vaccine production.

## Introduction

From research results, we know that elderly individuals with othercomorbidities such as hypertension, cardiovascular abnormalities, diabetes, cancer, and asthma are more severely affected with a higher rate of case mortality ^1-4^. Particularly, a higher prevalence of the disease is reported in individuals above 60 years of age ^1,5^. Besides, a retrospective study conducted on 85 fatal COVID-19 cases reported a median age of 65.8 years for diabetes, cardiopulmonary diseases, and hypertension comorbidities ^6^. Moreover, both innate and adaptive immune systems are reportedly weakened in the elderly with co-morbid, and the possibility of underlying chronic diseases upsurges as age advances, leading to the higher acquisition of infections ^7^. Studies have called attention to the age-related reduction in vaccine efficacy, owing to a decline in the adaptive and inherent immune response. As the aging population is increasing globally, vaccine endeavors must take into consideration age-related issues to ensure successful control of infectious diseases, like COVID-19. These attempts are necessary to secure a healthier life for the elderly as life expectancy is increasing ^8^. An effective vaccine must stimulate a broad T and B cell response, potentially overcoming the reduced immune function in the older population ^9^.

In the interim guidelines for COVID-19 vaccines from Pfizer, Moderna, and AstraZeneca published by the WHO Strategic Advisory Group of Experts on Immunization (SAGE), it was recommended that all the three COVID-19 vaccines can be used for individuals with high-risk comorbidities^10^.

In Bangladesh, from early February mass vaccination has started with AZD1222. In step by step mRNA1273, BBIBP-CorV started. In this study, we want to evaluate the effect of AZD1222, mRNA1273, BBIBP-CorV to produce antibodies in elderly people who have co-morbidities.

## Method

This observational study was conducted at Sheikh Hasina National Institute of Burn and Plastic Surgery, Dhaka February 2021 to October 2021. We enrolled 40 participants in each group and 120 are total number for our study age range between 60 years and above. Participants’ demographic data along with past SARS-COV-2 infection history and existing comorbidities were recorded using a structured questionnaire. Antibodies to the RBD of the S1 subunit of the viral spike protein (IgG) were performed from the plasma samples using a commercially available Anti-SARS-CoV-2 ELISA kit(DiaSino SARS-CoV-2 S1 RBD IgG Quant) as per instruction supplied by the manufacturer. The study protocol was reviewed and approved by the Institutional Review Committee of the Sheikh Hasina National Institute of Burnand Plastic Surgery, Dhaka (approval memo number: SHNIBPS/April-14(09)). Informed written consent was taken from each participant. Data were summarized by descriptive analysis. Numerical variables are reported as mean and standard deviation or median and interquartile ranges, as appropriate. Categorical variables are reported as counts and percentages. The analysis was performed in Graphpad Prism 9.0.

## Results

Among 120 participants with equal number of participants in each of the vaccine groups all of them were aged between 60-72 years, of whom 65% were males and 35 % were females. Among three vaccine groups, 100% of the mRNA-1273 recipients, 62.5% of AZD1222 recipients and only 2.5% of BBIBP-CorV recipients had injection site pain. Fever as AEFI was found among 32.5% of AZD1222 recipients and 10% of BBIBP-CorV recipients. None of the mRNA-1273 recipients had any fever as AEFI (Table 1).

**Table 1:**
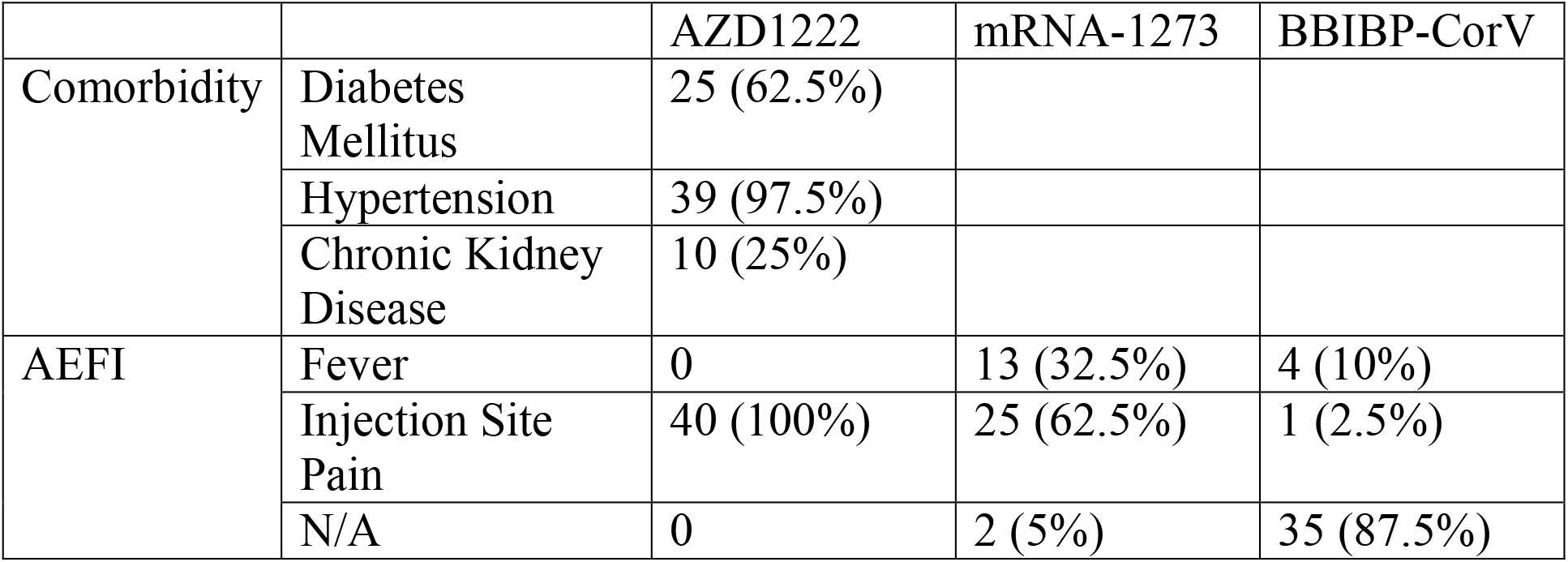
AEFI Characteristics among COVID-19 vaccine recipients with 14 days after receiving their 2^nd^ dose.

|  |  | AZD1222 | mRNA-1273 | BBIBP-CorV |
| --- | --- | --- | --- | --- |
| Comorbidity | Diabetes Mellitus | 25 (62.5%) |  |  |
|  | Hypertension | 39 (97.5%) |  |  |
|  | Chronic Kidney Disease | 10 (25%) |  |  |
| AEFI | Fever | 0 | 13 (32.5%) | 4 (10%) |
|  | Injection Site Pain | 40 (100%) | 25 (62.5%) | 1 (2.5%) |
|  | N/A | 0 | 2 (5%) | 35 (87.5%) |

Anti S1 RBD IgG was detected among all the participants from each vaccine groups after 14 days taking their 2^nd^ dose. A non-parametric multiple comparison test (Kruskal-Wallis test) of Anti S1 RBD IgG levels among three vaccine groups revealed significant differences (P-value <0.05) between groups (Figure 1). The IgG level was almost twice in mRNA-1273 group (mean 577.1± 44.33 AU/ml) compared to AZD1222(mean 308.5 ±37.91 AU/ml) and BBIBP-CorV group (mean 257.5 ±32.75 AU/ml).

**Figure 1:**
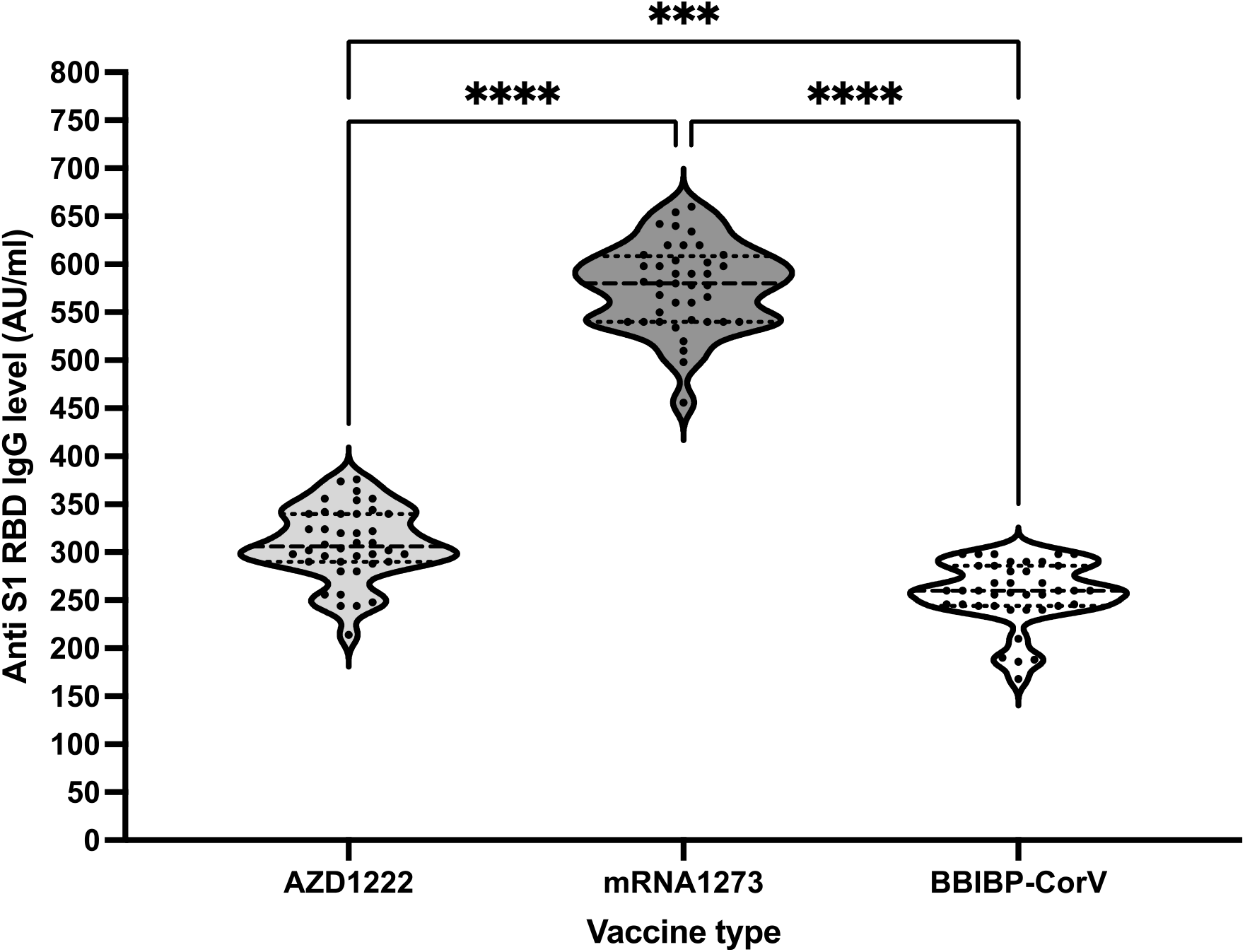
SARS-CoV-2 Anti S1 RBD IgG comparison among different COVID-19 vaccine recipients after completing both dosage

## Discussion

It is very difficult to interpret vaccine studies in the older population, due to the high heterogeneity observed with age and the multitude of underlying comorbidities found within the aging population. Cellular senescence, partial differentiation of naive T cells, reduced TCR signaling, causes reduced T-cell expansion and also altered functional of effector and/or memory T-cell responses lower antibody secretion by plasma cells that result in reduced levels of antibodies ^23^.

Vaccines designed to evoke protective immune responses endure the key hope for stopping the COVID-19 pandemic caused by SARS-CoV-2 10. Neutralizing antibodies are a likely correlate of protection against SARS-CoV-2 infection, as suggested by vaccine efficacy studies. From preclinical studies in mice and non-human primates, and data from the early use of convalescent plasma in elderly patients 11-12,17-20. There is increasing evidence that neutralizing responses are a correlate of protection ^11-13^.

mRNA-1273 (Moderna) (100μg mRNA, 2 doses, 28 days apart) mRNA-lipid nanoparticle encoding full-length S protein, modified by two proline mutations to lock protein in the pre-fusion conformation^14^. Antibody response in humans is S-binding antibody detected 14 days after the first dose, levels increased slightly by 28 days, with marked increase after the second dose^14^. Minimal Nab present after the first dose, peak at 14 days after the second dose^15^.

ChAdOx1 nCoV-19 (University of Oxford/ Astra-Zeneca) (2.5–5×1010 viral particles, 2 doses, ≥ 28days apart)^16^.Recombinant, replication-deficient simian adenovirus vector expressing the full-length S protein with a tPA leader sequence^17^. Antibody response in humans is: S-binding antibody present 14 days after the first dose, levels increased by 28 days17; marked increase after the second dose, peak at 14 days after the second dose; predominantly IgG3 and IgG1 ^18^; BBIBP-CorV (Sinopharm) (4µg protein, 2 doses, 21 days apart)^20^.SARS-CoV-2 grown in Vero cells, inactivated with β-propiolactone, and adsorbed onto aluminum hydroxide^19^.

Interferon-γ (IFNγ)-making T helper 1 cell (TH1 cells) are produced during acute infection, and it has been proposed that this T H1 cell-biased phenotype is linked with less severe disease^21,22^. Studies have proven that individuals with raised levels of IFNγ-secreting T cells against the S protein, nuclear proteins, and membrane proteins of SARS-CoV-2 may have better protection from disease^23^. Both mRNA and viral vector-based vaccines effectively produced TH1.

IgG subclasses are important in producing antibodies. From the different studies, we have found that mRNA vaccines induce all classes of IgG whereas viral vector-based vaccine especially induces IgG1 and IgG3.

Variability in somatic hyper-mutation could influence neutralization through antibody affinity maturation. The study has found that participants with elderly age have a low level of somatic hypermutation in class-switched B cell receptors (BCRs) than the younger group^10^. Neutralizing antibodies also waned overtime after the second dose of BNT162b2 or ChAdOx1^27^.

From our study, we have found that after getting two doses of vaccine mRNA produced a much higher antibody than viral vector and inactivated vaccine.

Whereas from AEFI data we have found that inactivated vaccines have fewer reports than others which is similar to published data ^24-26^.

### Limitations and Future Suggestions

This small sample size cannot produce generalizability. Therefore, subsequent clinical studies with larger sample sizes with different age group should be enclosed to confirm our findings. Moreover, only antibody response could not evaluate the efficacy of any vaccine, cellular response should be evaluated.

## Conclusion

COVID-19 vaccine is still produced by a few countries which can be counted by finger. A country like Bangladesh still depends on vaccine-producing countries and COVAX. Careful utilization of vaccines can be a possible way to prevent COVID-19 related death. Though our sample size is very small, from our study we can find that mRNA 1273 is best protective for the elderly population with different co-morbidity. Every vaccine is safe and effective whose is approved by WHO. Calculative use of the vaccine may produce outcomes for the future as we are still way behind in the proper amount of vaccine production.

## Data Availability

All data produced in the present work are contained in the manuscript

